# Sexual orientation and gender identity reporting in highly cited alcohol research

**DOI:** 10.1101/2023.03.29.23287850

**Authors:** Dean J. Connolly, Santino Coduri-Fulford, Connor Tugulu, Meron Yalew, Elizabeth Moss, Justin C. Yang

## Abstract

**Purpose:** This study aimed to measure the frequency of high-quality and transparent sexual orientation and gender identity data collection and reporting in highly cited alcohol use research, using the extant literature to identify community-informed priorities for the measurement of these variables.

**Methods:** A single search to identify alcohol use literature was conducted on PubMed with results restricted to primary research articles published between 2015 and 2022. The 200 most highly cited studies from each year were identified and their titles and abstracts reviewed against inclusion criteria following deduplication. Following full-text review and data extraction, the fidelity of the results was verified with a random sample prior to analyses.

**Results:** The final sample comprised 580 records. Few studies reported gender identity (n=19; 33.4%) and, of these, 7.2% reported the associated gender identity measure. A two-stage approach to measuring gender was adopted in five studies and 13 studies recorded non-binary gender identities (reported by 0.9% of the whole sample). Nineteen (3.3%) studies reported sexual orientation and more than half of these provided the sexual orientation measure. Five studies adopted a two-step approach to measuring gender identity and open offered a free-text response option. Eight of 20 studies reporting gender identity and sexual orientation measures were classified as sexual and gender minority specialist research.

**Conclusions:** Transparent and culturally competent gender identity and sexual orientation reporting is lacking in highly cited alcohol research.

## Introduction

A substantial body of research suggests that, relative to their majority counterparts, lesbian, gay, bisexual, transgender, queer and other sexual or gender minority (LGBTQ+) people experience a greater prevalence of high-risk or dependent drinking1,2 and are more likely to experience harms such as alcohol-related blackouts.^3^ This disparity is likely to widen as antecedents of problem alcohol use observed in this population, e.g., anti-LGBTQ+ legislation, hate crime and dehumanising media discourse, are increasing in prevalence and severity internationally.

Despite the high acceptability, among both the general population and LGBTQ+ communities, of sexual orientation and gender identity (SOGI) data collection,^4^ SOGI is rarely recorded in electronic health records, health data systems and large epidemiological surveys,^5^ resulting in a paucity of quantitative data. Inclusion in these datasets is necessary to monitor trends in drinking, the scale and correlates of harm as well as engagement with and response to healthcare. A recent United Kingdom National Institute for Health Research review identified no randomised controlled trials (RCTs) which collected SOGI data, suggesting this disparity is also an issue in experimental research.

Alcohol use research with LGBTQ+ people has largely involved within-group studies which recruit small convenience samples, frequently comprised of people with an additional shared vulnerability, e.g., sex work, primarily aimed at HIV risk reduction or understanding the relationship between LGBTQ+ status and alcohol harm (specialist research).^2^ The extent to which sexual or gender minority status are recorded or used in general population alcohol research and how this practice compared to specialist research is unclear and bears further scrutiny.

Existing studies with LGBTQ+ samples have identified several key characteristics which define good SOGI data collection and reporting practice.7–11 The first relates to the inclusion of non-binary/gender diverse people in data collection and reporting.^11^ While the data collected with this approach is unlikely to generate sufficient data for stratified analysis, non-binary/gender diverse participants should still be identified and their data summarised and reported with a view to informing future meta-analyses.

The literature overwhelmingly supports the use of a two-stage approach (i.e., asking gender identity and birth-registered sex, in either order) to understand both gender identity and trans status. Quantitative and qualitative work provide evidence of high sensitivity and specificity with this method, which is largely accepted within the LGBTQ+ community.8,11 An open-ended response option when measuring SOGI was generally endorsed in the extant literature,^9,11^ with one study participant advising “put a line and let [us] put what [we] want [our] damn self”, highlighting the potential for identity invalidation with aggregated response categories.^9,^

This aim of this study was to measure the frequency of high-quality and transparent SOGI data collection and reporting in highly cited alcohol use literature. A secondary aim was to identify patterns in current practice with measures described above as indicators of good practice.

## Methods

Alcohol was selected as the sole substance of interest because it is the most ubiquitously used intoxicant globally,^12^ and there is a substantial and growing body of specialist LGBTQ+ alcohol literature.2,13 Highly cited literature was investigated for two reasons. The first related to the observation that specialist literature was cited infrequently, relative to the whole sample. By investigating highly cited research, we were able to address the risk that specialist research in the sample might inflate estimates of SOGI representation. Secondly, LGBTQ+ people deserve to be represented in and benefit from the most impactful research, as a matter of health equity. If this is not currently the case, it must be highlighted and addressed.

This article presents a secondary analysis of SOGI recording and reporting in highly cited alcohol use research. Preferred Reporting Items for Systematic Reviews and Meta-Analyses (PRISMA) informed the sampling (i.e., search, title-abstract screening) and data extraction.

### Search strategy

A single search was conducted on PubMed on 31st May 2022. MeSH terms “alcoholism” (MeSH ID: D000437) and “binge drinking” (MeSH ID: D063425) were combined with key words “alcohol use disorder”, “alcohol consumption”, and “alcohol dependence”, with the Boolean classifier “OR”. Results were then restricted to years 2015-2022, inclusive, and the following article type filters applied: Clinical Study, Clinical Trial, Clinical Trial, Phase III, Clinical Trial, Phase IV, Comparative Study, Controlled Clinical Trial, Multicenter Study, Observational Study, Pragmatic Clinical Trial, Randomized Controlled Trial, Twin Study, Validation Study, Humans.

Bibliographic data from all records identified by the search were downloaded to Zotero, a reference management software.^15^ A software ‘add-on’, Zotero Citation Counts Manager 1.3.0, was applied to extract, from Crossref, the number of times each record had been cited.^16^ The reliability of the citation count was tested using a random sample (n=30) from a pilot search. The 200 most highly cited records from each year were retained and collated to give a study sampling frame of 1,600 records.

### Inclusion and exclusion criteria

With no restriction by study design, all original qualitative, quantitative, or mixed-methods studies, with more than ten human participants, and at least one alcohol use variable were included. Research with non-human subjects and all forms of journal communication not classified as original research, including (systematic) reviews, case reports and series, were excluded. Conference proceedings, books and chapters were also excluded.

### Record selection and data extraction

Bibliographic data for the entire sample were uploaded to Rayyan, a systematic review software, which has a partially automated de-duplication function.^17^ All authors were given access to the database and a minimum of two independently assessed each abstract against the inclusion criteria. Conflicts were resolved by the first author who made the final decision regarding inclusion. Included records were then divided equally among all authors. A study’s eligibility was confirmed by a review of the full text and a piloted data extraction table was populated with data from each study by at least one author. These data included study characteristics (DOI, authors, year of publication) and a list of pre-specified criteria suggesting high-or low-quality SOGI data collection. These criteria began with determination of whether SOGI was measured (with SOGI measures collected verbatim, if available). Also assessed was the use of a two-stage approach to measuring gender identity and whether an open-ended response option was provided for both gender identity and sexual orientation measures. The recording and reporting of non-binary/gender diverse participants’ data was assessed, as was the frequency of two sets of response options to measure gender identity which represent poor practice: 1. male, female, transgender; 2. male, female, prefer not to say. The operationalisation of each variable was reviewed with and agreed by the whole team to ensure consistency.

### Analysis

A random sample (5%) of the data was reviewed by the first author to confirm fidelity. The number of records fulfilling each pre-specified outcome was described as a count and percentage of eligible records, e.g., the number of articles reporting a gender identity measure was reported as a percentage of those which reported participants’ gender identity. Verbatim SOGI measures were tabulated.

## Results

### Search results

A total of 29,096 records were identified from 2015 (n=3,737), 2016 (n=3,710), 20 (n=3,939), 2018 (n=3,918), 2019 (n=3,883), 2020 (n=4,156), 2021 (n=4,210) and 20 (n=1,543). Following deduplication (N=1,450), title-abstract screening (N=621), full-text review and retrieval (N=581) and exclusion of one retracted article, the final sample comprised N=580 (Figure 1).

**Fig 1.**
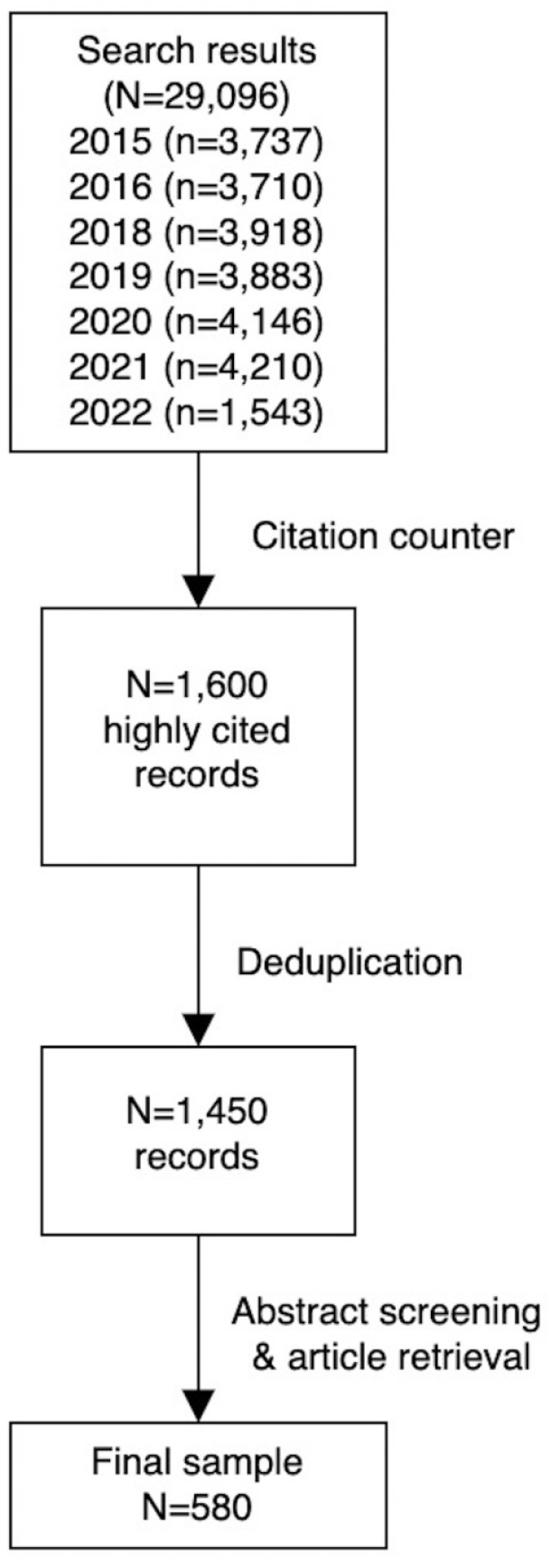
Flowchart of record selection.

### SOGI recording and reporting practices

One hundred and ninety-four studies (33.4%) reported participants’ gender identity. Of these, 14 (7.2%) reported the gender identity measure (e.g., the question/query/prompt and response options given to study participants; Table 1).^18–31^ Five studies (35.7%; 0.9% of whole sample) adopted a two-stage approach,^25–28,31^ and one used an open-ended question with a free-text response option.

**Table 1:** Gender identity measures in highly cited alcohol research published 2015-2022

| Authors (year) | Gender identity measure (i.e., question or stem) and response options |
| --- | --- |
| Auerbach et al. (2018, 2019),<sup>19,20</sup><br>Ebert et al. (2019)<sup>18</sup> | Gender was assessed by asking respondents whether they identified as being male, female, transgender (male-to-female/female-to-male), or “other” |
| Agley & Xiao (2021)<sup>21</sup> | Q: Please indicate your gender<br>R: male, female, non-binary, transgender |
| Agley et al. (2021)<sup>22</sup> | Q: Please indicate your gender<br>R: female, male, transgender, other |
| Callinan et al. (2021)<sup>23</sup> | Participants were asked for their “self-identified gender”<br>R: male, female, non-binary, not-listed gender |
| Kilian et al. (2021)<sup>24</sup> | Q: please specify your gender<br>R: male, female, other |
| Coulter et al. (2018)<sup>25</sup> | Q1 (to ascertain LGBT status): which of the following best describes you? Mark all that apply<br>R1: heterosexual (straight), gay or lesbian or bisexual, transgender, not sure (*removed from analysis*), decline to respond (*removed from analysis*)<br>Researchers “created a three-category measure of gender by coding participants who selected “transgender” as transgender adolescents, and those who did not select this option were coded as cisgender males or cisgender females based on their responses to the following question”<br>Q2: what is your sex?<br>R2: male, female |
| Davies et al. (2022)<sup>26</sup> | Q1: What is your gender?<br>R1: male, female, non-binary, different identity<br>Q2: What gender were you assigned at birth?<br>R2: male, female |
| Every-Palmer et al. (2020)<sup>27</sup> | Q1: which gender do you identify with?<br>R1: male, female, gender diverse<br>Q2: are you?<br>R2: transgender female to male, transgender male to female, intersexed, gender non-conforming, genderqueer, two-spirit, third gender |
| Hegazi et al. (2016)<sup>28</sup> | “gender identity...stated by the PERSON” (during clinical contact/appointment booking etc.)<br>R: man (including trans man), woman (including trans woman), non-binary, other (not listed), not stated |
| Li et al. (2020)<sup>29</sup> | Q: Your gender:<br>R: *free-text option only*<sup>a</sup> |
| Thompson et al. (2021)<sup>30</sup> | Q: what is your gender identity?<br>R: man, woman, non-binary |
| Reisner et al. (2016)<sup>31</sup> | *Q not reported* (“eligible participants were assigned male sex at birth”)<br>R: woman, female, transgender woman, transfemale, male-to-female, other identity on the transfeminine spectrum |

**Notes:** <sup>a</sup>: though only data for males and females included in report. The exclusion of transgender people was acknowledged as a limitation; LGBT: lesbian, gay, bisexual, transgender; Q: question/query; R: response

Thirteen studies (6.7% of those reporting gender identity) recorded non-binary identities^21,23,24,26–28,30–36^ and, of these, five (38.5%; 0.9% of whole sample) reported non-binary participants’ data.^26,31–34^ Five studies listed ‘male’, ‘female’ or ‘transgender’ as mutually exclusive response options.^18–20,22,25^ One study gave ‘male’, ‘female’, or ‘prefer not to say’ as mutually exclusive response options.^37^ Sexual orientation was reported in studies (3.3%). Eleven of these (57.9%) reported the sexual orientation measure (Table 2).^18– 20,25,28,38–43^ Of the 20 unique studies reporting SOGI measures,^18–25,27–31,38–43^ eight studies (40.0%) were SOGI specialist research.^25,28,31,38,41–43^ There was no additional SOGI specialist research in the wider sample.

**Table 2:**
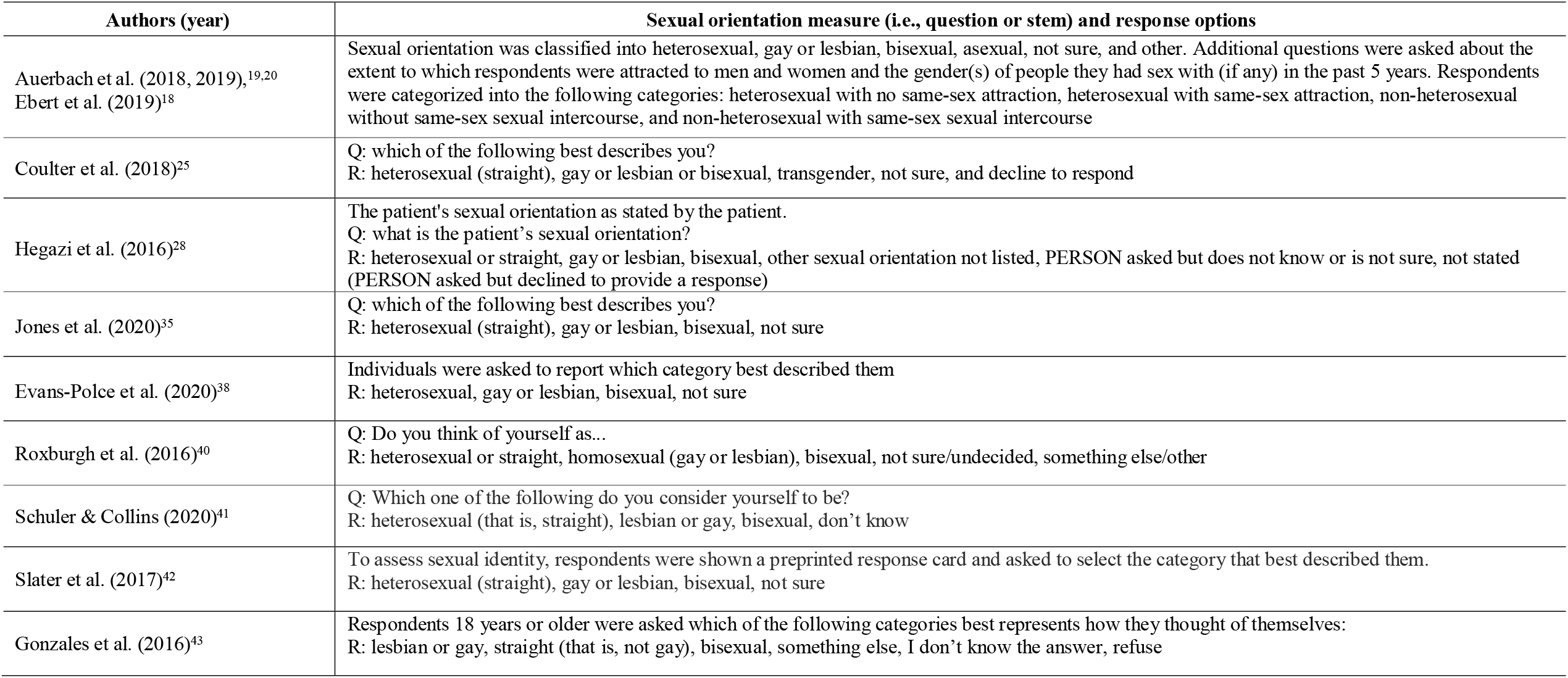
Sexual orientation measures in highly cited alcohol research published 2015-2022

| Authors (year) | Sexual orientation measure (i.e., question or stem) and response options |
| --- | --- |
| Auerbach et al. (2018, 2019),<sup>19,20</sup><br>Ebert et al. (2019)<sup>18</sup> | Sexual orientation was classified into heterosexual, gay or lesbian, bisexual, asexual, not sure, and other. Additional questions were asked about the extent to which respondents were attracted to men and women and the gender(s) of people they had sex with (if any) in the past 5 years. Respondents were categorized into the following categories: heterosexual with no same-sex attraction, heterosexual with same-sex attraction, non-heterosexual without same-sex sexual intercourse, and non-heterosexual with same-sex sexual intercourse |
| Coulter et al. (2018)<sup>25</sup> | Q: which of the following best describes you?<br>R: heterosexual (straight), gay or lesbian or bisexual, transgender, not sure, and decline to respond |
| Hegazi et al. (2016)<sup>28</sup> | The patient's sexual orientation as stated by the patient.<br>Q: what is the patient's sexual orientation?<br>R: heterosexual or straight, gay or lesbian, bisexual, other sexual orientation not listed, PERSON asked but does not know or is not sure, not stated (PERSON asked but declined to provide a response) |
| Jones et al. (2020)<sup>35</sup> | Q: which of the following best describes you?<br>R: heterosexual (straight), gay or lesbian, bisexual, not sure |
| Evans-Polce et al. (2020)<sup>38</sup> | Individuals were asked to report which category best described them<br>R: heterosexual, gay or lesbian, bisexual, not sure |
| Roxburgh et al. (2016)<sup>40</sup> | Q: Do you think of yourself as...<br>R: heterosexual or straight, homosexual (gay or lesbian), bisexual, not sure/undecided, something else/other |
| Schuler & Collins (2020)<sup>41</sup> | Q: Which one of the following do you consider yourself to be?<br>R: heterosexual (that is, straight), lesbian or gay, bisexual, don't know |
| Slater et al. (2017)<sup>42</sup> | To assess sexual identity, respondents were shown a preprinted response card and asked to select the category that best described them.<br>R: heterosexual (straight), gay or lesbian, bisexual, not sure |
| Gonzales et al. (2016)<sup>43</sup> | Respondents 18 years or older were asked which of the following categories best represents how they thought of themselves:<br>R: lesbian or gay, straight (that is, not gay), bisexual, something else, I don't know the answer, refuse |

## Discussion

### Summary of key findings

One third of studies reported participants’ gender identity. However, markedly fewer (<1%) reported the associated measure with a high proportion indicating a poor understanding of gender identity. Only one used an open-ended gender identity measure. Non-binary/gender diverse participants’ data was recorded in a small minority of studies. Less than half of these reported stratified analyses of non-binary/gender diverse participants’ data. While fewer studies reported sexual orientation, a greater proportion of sexual orientation than gender identity measures were reported. Almost half of the studies reporting a SOGI measure were classified as specialist research.

### Findings in context

Corroborating the findings presented here, a 2015 study similarly identified poor representation of LGBTQ+ people in alcohol research. Examining research published in 20 and 2012, the authors found that sexual orientation was reported in 2.3% (PsycINFO) and 6.4% (PubMed) of sampled “substance abuse” articles from 2012.^44^ Non-binary gender was reported in 2.3% (PsycINFO) and 1.9% (PubMed) of the same sample. The authors observed a negligible improvement from 2007.^44^ Comparing these results with the findings of the present (2015-2022) study, it appears that SOGI data reporting and LGBTQ+ representation in alcohol research has declined over time.^44^ However, the variables measured in the two studies are not directly comparable and the narrow selection of RCTs in 2016, where investigators have greater control over which data they collect, may have inflated earlier estimates of good practice.^44^ Moreover, it is possible that a larger sample was required to map over such short time periods.

Poor recording and reporting of ethnicity has also been observed. A recent systematic review examining RCTs of pharmacotherapies for alcohol use disorder found that 49.0% of included records had not reported their participants’ ethnicity.^45^ While the difference in population size and the circumstances of their exclusion preclude direct comparison, it appears both these minoritised groups are underrepresented in alcohol research.

### Strengths and limitations

Strengths of this study include its partial adherence to PRISMA e.g., double, independent title-abstract review, pre-specified inclusion/exclusion criteria and study variables, and piloted data extraction with accuracy checking.^14^ Limited resources precluded double screening of full-texts and whole sample double data extraction.

Conflation between gender and sex in the primary literature meant it was frequently difficult to determine whether participants’ gender identity had been reported. Interchangeable use of terms “sex” and “gender”, “female” and “woman”, “male” and “man” between and within articles may have resulted in misestimation of the frequency of gender identity reporting. These findings may not be generalisable to the wider alcohol use literature as only the most highly cited studies from one database were sought and included. However, inclusion in the most impactful research is a matter of health equity. Minority groups are entitled to be represented in studies which are more readily translated into public, group, or individual health interventions.

### Implications for policy, research, and practice

Inclusion of community- and expert-informed SOGI measures in all observational and experimental research should be enforced by grantors and research ethics committees. Healthcare providers should be supported to adapt their electronic records to collect SOGI data with cultural competence being mindful that approaches will likely evolve with time.

The exclusion of LGBTQ+ people from alcohol research, through inadequate recruitment, data collection or statistical stratification may mean that caution is required when administering interventions in the absence of valid outcome (favourable or adverse) data. A person-centred approach to supporting alcohol service users is required.

Despite LGBTQ+ people reporting dissatisfaction with the use of broad catch-all response options to supposed gender identity measures e.g., ‘transgender’, these are not uncommon. In future, researchers should consider using a qualitative measure to empower participants to disclose their exact gender identity. Code to categorise these data for analysis has been trialled with success.

## Conclusion

Transparent and culturally competent SOGI reporting is lacking in highly cited alcohol research. Alcohol researchers must comprehensively assess and document SOGI to fully understand and appropriately respond to the disproportionate alcohol-related harm experienced in LGBTQ+ communities.

## Data Availability

All data produced in the present study are available upon reasonable request to the authors

## Acknowledgments

None.

## Author Contributions

**Dean Connolly:** Conceptualisation (lead), Methodology (lead), Formal analysis (lead),

Investigation (lead), Writing – Original Draft (lead), Writing – Review & Editing (lead), Project administration (lead), Supervision (lead) **Santino Coduri-Fulford:** Investigation (equal), Writing – Review & Editing (equal) **Connor Tugulu:** Investigation (equal), Writing – Review & Editing (equal) **Meron Yalew:** Investigation (equal), Writing – Review & Editing (equal) **Elizabeth Moss:** Investigation (equal), Writing – Review & Editing (equal) **Justin Yang:** Methodology (equal), Investigation (equal), Writing – Review & Editing (lead)

## Disclaimer

None.

## Author disclosures

The authors have no conflicts of interest to declare.

## Funding

This research did not receive any specific grant from funding agencies in the public, commercial or not-for-profit sectors.

## Notes

### Competing Interest Statement

The authors have declared no competing interest.

### Funding Statement

This study did not receive any funding

